# COVID-19 infection and subsequent psychiatric morbidity, sleep problems and fatigue: analysis of an English primary care cohort of 226,521 positive patients

**DOI:** 10.1101/2021.06.24.21259463

**Authors:** Kathryn M Abel, Matthew J. Carr, Darren M. Ashcroft, Trudie Chalder, Carolyn A. Chew-Graham, Holly Hope, Navneet Kapur, Sally McManus, Sarah Steeg, Roger T. Webb, Matthias Pierce

**Affiliations:** Greater Manchester Mental Health Trust, Manchester, UK; Centre for Women’s Mental Health, University of Manchester, Manchester, UK; Division of Psychology and Mental Health; Centre for Pharmacoepidemiology and Drug Safety; National Institute for Health Research Greater Manchester Patient Safety Translational Research Centre; Manchester Academic Health Science Centre, Manchester, UK; Department of Psychological Medicine, Institute of Psychiatry, Psychology & Neuroscience, King’s College London, London, UK; School of Medicine, Keele University, Keele, UK; National Centre for Social Research, London, UK; School of Health Sciences, City, University of London, London, UK; Faculty of Biology, Medicine and Health Sciences, Manchester, UK

**Author notes:** Corresponding author:* | Address Room 3.307 Jean McFarlane Building University of Manchester, Oxford Road, Manchester. M13 9P |.

## Abstract

**Objectives:** The primary hypothesis was that the risk of incident or repeat psychiatric illness, fatigue and sleep problems increased following COVID-19 infection. The analysis plan was pre-registered (https://osf.io/n2k34/).

**Design:** Matched cohorts were assembled using a UK primary care registry (the CPRD-Aurum database). Patients were followed-up for up to 10 months, from 1^st^ February 2020 to 9^th^ December 2020.

**Setting:** Primary care database of 11,923,499 adults (≥16 years).

**Participants:** From 232,780 adults with a positive COVID-19 test (after excluding those with <2 years historical data or <1 week follow-up), 86,922 without prior mental illness, 19,020 with anxiety or depression, 1,036 with psychosis, 4,152 with fatigue and 4,539 with sleep problems were matched to up to four controls based on gender, general practice and year of birth. A negative control used patients who tested negative for COVID-19 and patients negative for COVID with an influenza diagnosis.

**Main Outcomes and Measures:** Cox proportional hazard models estimated the association between a COVID-19 positive test and subsequent psychiatric morbidity (depression, anxiety, psychosis, or self-harm), sleep problems, fatigue or psychotropic prescribing. Models adjusted for comorbidities, ethnicity, smoking and BMI.

**Results:** After adjusting for observed confounders, there was an association between testing positive for COVID-19 and almost all markers of psychiatric morbidity, fatigue and sleep problems. The adjusted hazard ratio (aHR) for incident psychiatric morbidity was 1.75 (95% CI 1.56-1.96). However, there was a similar risk of incident psychiatric morbidity for those with a negative COVID-19 test (aHR 1.57, 95% CI 1.51-1.63) and a larger increase associated with influenza (aHR 2.97, 95% CI 1.36-6.48).

**Conclusions:** There is consistent evidence that COVID-19 infection elevates risk of fatigue and sleep problems, however the results from the negative control analysis suggests that residual confounding may be responsible for at least some of the association between COVID-19 and psychiatric morbidity.

## Introduction

Many people infected with SARS-CoV-2 (COVID-19) experience symptoms beyond the acute phase of the illness, particularly fatigue, ‘brain fog’ and sleep problems[1,2]. There is also concern that infection is causing mental illness and mechanisms linking the immune system, inflammation and the brain have been proposed[3,4]. Two registry studies using the same North American database reported an increased risk of a range of mental illnesses (including anxiety, depression and psychosis) associated with confirmed COVID-19 (regardless of hospitalisation) compared to controls with other viral infections[5,6]. In contrast, a Danish registry study compared people with non-hospitalised COVID-19 infection with people who tested negative and did not find an association with anxiety or depression[7]. These divergent findings may be because different choices of exposures and controls may lead to different confounding structures.

Registry studies that seek to quantify the effect of COVID-19 infection may be confounded by several sources affecting the likelihood that somebody is infected (e.g. their occupation), the likelihood they present to services (e.g. comorbidities), or the likelihood they receive a test (e.g. health anxiety). For example, in the UK, only 25% of people with COVID-19 symptoms present for testing[8] and it is likely that those who do form a selective group.

When unobserved confounding is suspected, the validity of causal claims can be examined using a ‘negative control’[9]. Here, a variable, with no conceivable effect on the outcome, but with a similar confounding structure, is substituted for the exposure under investigation. If the result from the negative control is similar to that observed using the primary exposure, then unobserved confounding is implicated.

We examined the effect of COVID-19 infection on indicators of psychiatric disorder (anxiety, depression, self-harm, psychosis and prescription of psychotropic medication), sleep problems and fatigue using UK primary care data. In separate matched cohorts, we consider both incident events as well as further events for people with pre-existing mental illness, fatigue or sleep problems. Our primary hypothesis was that COVID-19 infection increased the likelihood of new or repeat presentation of psychiatric morbidity, sleep problems or fatigue symptoms. We also hypothesised that the increase in risk would be greatest for women and those living in more deprived areas. We used a negative control design that include a cohort with a negative test. Finally, we investigated the specificity of infection with COVID-19 and psychological outcomes by repeating our analysis in individuals with a flu diagnosis.

## Methods

This paper is prepared in line with the RECORD statement[10]. The analysis plan was agreed with all authors prior to analysis and is available online (https://osf.io/n2k34/).

### Data source

For this retrospective study, matched cohorts were assembled from the Clinical Practice Research Datalink (CPRD-Aurum) dataset: a large UK primary care registry covering 19 million patients[11]. It contains information on clinical events recorded by healthcare professionals, including diagnosis, symptoms and therapies.

### Eligible cohort

Eligible patients were those registered at a CPRD-Aurum participating clinical practice 1^st^ February to 8^th^ December 2020 and aged 16 or over during 2020. Eligible follow-up began on the latest date of: 1^st^ February 2020, or their clinical practice registration, and ended at the earliest date of: their death, the date they transferred out of a clinical practice, or the end of data collection (9^th^ December 2020). Patients were excluded if they had an indeterminate gender recorded (total n=394). This resulted in 11,923,105 patients for matching, 84% of whom had the maximum 44.6 weeks of follow-up.

### Exposure

COVID-19 infected patients included people with a positive polymerase chain reaction (PCR) test. The clinical codes used to select cases were developed by the UK’s Medicines and Healthcare Products Regulatory Agency[12]. In the UK, whilst the majority of testing took place in the community, in primary care patients presenting with symptoms consistent with COVID-19 infection were PCR-tested. In addition, since 20^th^ July 2020 doctors in primary care were notified of all PCR test results, regardless of outcome[13]. In total, 232,780 out of 11,923,105 eligible patients (2.0%) had a record of a positive COVID-19 test during the observation period.

### Outcome

The outcomes included a diagnosis with or symptoms of; depression, anxiety disorders, self-harm (including self-poisoning and self-injury episodes of varying suicidal intent), affective/non-affective psychosis, sleep problems, and fatigue or fatigue-like syndromes (e.g. post-viral fatigue syndrome). Additionally, outcomes included prescriptions for; antidepressants, anxiolytics, antipsychotics, mood stabilisers; benzodiazepines; and non-benzodiazepines hypnotics. There were too few cases of post-traumatic stress disorder (PTSD) in the incident matched cohort to enable separate examination of these diagnoses so, in a departure from our protocol, these were combined with anxiety disorders. Outcome codes in this analysis were identified in a prior analysis and are published online (see https://clinicalcodes.rss.mhs.man.ac.uk/medcodes/article/173/). Two senior clinical academics (CAC-G, NK) and a senior academic pharmacist (DMA) reviewed the clinical and medication code lists, respectively.

### Covariates

Covariates were identified from 10 years of linked patient records. Smoking status (current, former, never) and body mass index (BMI) were identified from the latest available data. Comorbidities were identified using indicator of diseases used to construct the Charlson comorbidity index[14]: cancer, cerebrovascular disease, chronic pulmonary disease, congestive heart failure, connective tissue disease, dementia, diabetes (with/without end organ damage), HIV/AIDS, hemiplegia, myocardial infarction (MI), liver disease, renal disease, peripheral vascular disease, posterior vitreous detachment (PVD), and peptic ulcer disease. Additional data were extracted on gender, year of birth, ethnicity (White, Asian, Black, Mixed, other) and the practice’s Index of Multiple Deprivation (IMD) score, an area-level ranking of socioeconomic deprivation divided into quintiles according to the national distribution. Additional variables captured where a doctor suspected COVID-19 infection, or where a patient presented for testing but received a negative test result.

### Matched cohorts

The analysis consisted of five matched cohorts by matching those with a positive COVID-19 test over follow-up with unexposed controls (Figure 1). Patients’ earliest date of positive diagnosis defined the ‘index date’, used to apply matching criteria and define the start of follow-up. The first cohort (termed ‘incident cohort’) excluded patients with recorded histories of mental illness, mental illness symptoms, self-harm, fatigue, sleep problems or psychotropic medications in the five years prior to their index date. The remaining four matched cohorts comprised people with a recent history of common mental illness, psychosis, sleep problems, or fatigue. These were defined using records from the six months prior to the index date. For both common mental illness and psychosis cohorts, an additional criterion was that patients had been prescribed antidepressants/anxiolytics or antipsychotics/mood stabilisers, respectively, in the last six months.

**Figure 1:**
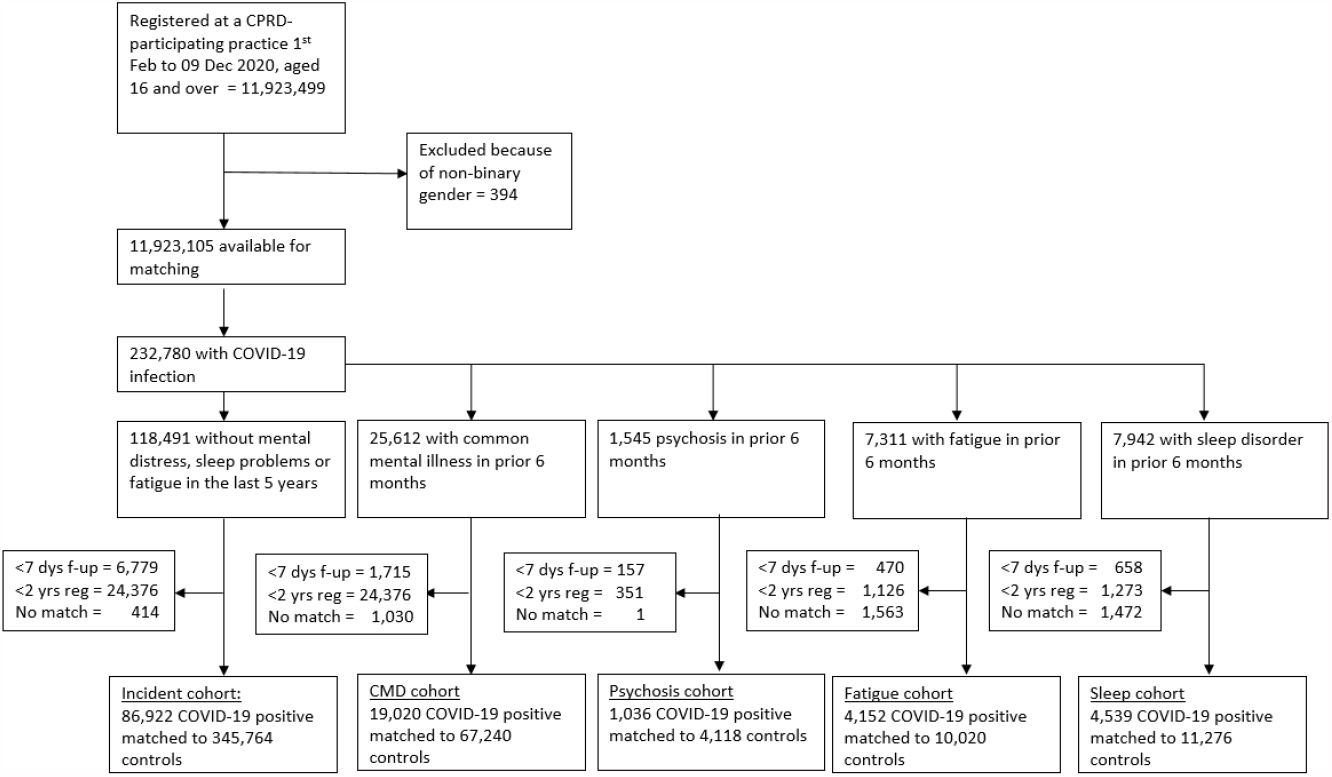
flow chart showing selection in to matched cohorts

Exclusions included patients with less than one week of eligible follow-up from their index date or less than two years registration prior to the index date. Patients were ineligible for the control if they had a record indicating suspected or confirmed COVID-19 in their history or in the week following the index date. After these criteria were applied, up to four control patients were selected for each exposed patient, regardless of whether or not they had been selected previously (i.e. matching with replacement). Cohorts matched on gender, clinical practice and year of birth except for the psychosis cohort (where there were fewer available unexposed comparator patients), which matched on gender and practice.

### Statistical analysis

Censoring occurred at the earliest date of: an outcome event (within each category examined), death, when the practice ceased collecting data, or the patient transferred to another practice. Hazard ratios were estimated using Cox proportional hazard models, stratified on each matched set. Interactions included a COVID-19 positive test result and; age (categorised as 16-24, 25-34, 35-49, 50-59, 60-69, 70-79, 80+), gender, Index of Multiple Deprivation (IMD) quintile of the general practice, and follow-up time elapsed since index date (<1 month, 1-<3 months, 3-<6 months or 6-10 months).

Adjusted models included ethnicity, comorbidities, smoking status, and BMI. To account for missing BMI, ethnicity and smoking information datasets were imputed ten times using chained equations and models included all covariates, a variable indicating the size of the general practice, and the patient’s height or weight. Each imputed dataset was fitted with a model and estimates combined using Rubin’s rules.

### Additional cohorts

Two further cohorts were constructed. The first substituted those with negative COVID-19 tests for those with a positive COVID-19 test (N = 560,495 negative test cases matched with 2,232,733 comparators) and proceeded with the same analysis. The second repeated the analysis using those recorded by their GP as having influenza or flu-like symptoms and with a negative COVID-19 test within two weeks. For the negative test cohort, we expected that there would be little (or no) effect of receiving a negative test (vs. comparators). Any divergence from a null finding would indicate potential bias from unobserved confounders. Notable potential confounders missing from the dataset include occupation, urbanicity and health-related anxiety. For the flu cohort, we expected to see an increase in fatigue[15], but a smaller effect than that seen for COVID-19[5].

### Sensitivity analysis

Two pre-planned sensitivity analyses were conducted. For the first, we investigated whether exposed patients identified after the introduction of widespread testing were different to those identified before by fitting an interaction between period (pre 1^st^ September 2020 or post) and the exposure. The final sensitivity analysis, a propensity score was calculated and used in the adjusted analysis, instead of adjusting for individual covariates. For calculation of the propensity score, missing data items were given separate categories and interactions between variables were included. Stata (version 16) was used for all analyses and graphs produced using the ggplot package in R.

### Patient involvement

This research utilised secondary data; the design, analysis and writing of the manuscript did not involve patient involvement.

## Results

Of 11,923,105 patients in the eligible cohort, 232,780 (2.0%) were recorded as having a positive PCR test for COVID-19 during their follow-up. The majority of positive tests occurred during two periods: the first in April and May 2020 and the second in October and November 2020 (see Appendix Supplementary Figure 1). Individuals with a positive test were more likely than those without to be female (56.2% vs 50.3%, Table 1) and in the youngest or oldest age groups (16-24 years 19.5% vs 14.3%; 80+ years 6.9% vs 5.9%). Those with a positive COVID-19 test tended to have higher BMI (median, IQR: 26.4, 22.9-30.8 vs 25.8, 22.6-29.7) and more recorded comorbidities (22.1% vs. 19.7% with 1 or more co-morbid condition). They were also more likely to have a clinical record in the preceding five years signifying psychiatric morbidity, fatigue, sleep problems or a psychotropic medication prescription.

**Table 1:** Description of available cohort according to whether they tested positive for COVID-19 over follow-up (prior to matching)

| Statistic | COVID-19 positive, n=232,780,<br>N (%) | Not positive, n=11,690,325<br>N (%) |
| --- | --- | --- |
| Gender |  |  |
| Female | 130,775 (56.2) | 5,880,245 (50.3) |
| Male | 102,005 (43.8) | 5,810,080 (49.7) |
| Age group |  |  |
| 16-24 | 45,456 (19.5) | 1,665,762 (14.3) |
| 25-34 | 41,336 (17.8) | 2,208,954 (18.9) |
| 35-49 | 56,983 (24.5) | 2,881,606 (24.7) |
| 50-59 | 39,119 (16.8) | 1,827,489 (15.6) |
| 60-69 | 21,359 ( 9.2) | 1,362,308 (11.7) |
| 70-79 | 12,444 ( 5.4) | 1,060,213 ( 9.1) |
| 80+ | 16,083 ( 6.9) | 683,993 ( 5.9) |
| Ethnicity |  |  |
| White | 152,885 (79.5) | 7,545,915 (80.3) |
| Asian | 25,988 (13.5) | 996,746 (10.6) |
| Black | 7,540 ( 3.9) | 499,926 ( 5.3) |
| Mixed | 3,325 ( 1.7) | 185,221 ( 2.0) |
| Other | 2,646 ( 1.4) | 175,694 ( 1.9) |
| Missing | 40,396 ( * ) | 2,286,823 ( * ) |
| BMI category |  |  |
| Underweight (<18.5) | 7,964 ( 3.9) | 386,194 ( 3.9) |
| Normal (≥18.5 & <25) | 73,790 (36.1) | 3,975,991 (39.9) |
| Overweight (≥25 & <30) | 64,222 (31.4) | 3,217,104 (32.3) |
| Obese (≥30 & <40) | 49,321 (24.1) | 2,058,755 (20.7) |
| Very obese (≥40) | 9,106 ( 4.5) | 323,657 ( 3.3) |
| Missing | 28,377 ( * ) | 1,728,624 ( * ) |
| Median years at clinical practice [IQR] | 9.6 [2.3 – 21.0] | 9.4 [2.7 – 21.4] |
| Number of comorbidities<sup>a</sup> |  |  |
| None | 181,280 (77.9) | 9,386,867 (80.3) |
| 1 | 26,143 (11.2) | 1,332,044 (11.4) |
| 2-4 | 23,262 (10.0) | 922,004 ( 7.9) |
| 5+ | 2,095 ( 0.9) | 49,410 ( 0.4) |
| Psychiatric morbidity in the last 5 years<sup>***</sup> |  |  |
| Depression | 38,727 (16.6) | 1,601,431 (13.7) |
| Anxiety | 31,072 (13.4) | 1,271,468 (10.9) |
| Psychosis | 2,941 ( 1.3) | 136,001 (1.2) |
| Eating disorder | 1,344 ( 0.6) | 47,018 (0.4) |
| Personality disorder | 726 ( 0.3) | 42,531 (0.4) |
| Self-harm | 2,975 ( 1.3) | 137,826 (1.2) |
| Fatigue | 18,396 ( 7.9) | 665,652 (5.7) |
| Sleep disorder | 16,759 ( 7.2) | 669,528 (5.7) |
| Medication in the last 5 years<sup>b</sup> |  |  |
| Antidepressants | 65,364 (28.1) | 2,681,383 (22.9) |
| Benzodiazepines | 20,571 ( 8.8) | 826,794 ( 7.1) |
| Nonbenzodiazepine hypnotics | 12,596 ( 5.4) | 542,565 ( 4.6) |
| Antipsychotics | 5,228 ( 2.3) | 209,463 ( 1.8) |
| Mood stabilisers | 15,306 ( 6.6) | 586,523 ( 5.0) |

\*Missing data excluded from percentages; <sup>a</sup>List of comorbidities from the Charlson comorbidity index; <sup>b</sup>From date of infection for COVID-19 positive group, from start of follow-up for the unexposed group

### Outcomes in those without prior mental illness, fatigue or sleep problems

The median follow-up of the incident-matched cohort was 6.3 weeks (interquartile range: 4.0-9.3 weeks). After matching on age, gender and registered practice, and adjusting for ethnicity, smoking status, BMI, and comorbidities, there was a 75% increase in the risk of any psychiatric morbidity (adjusted HR(aHR): 1.75, 95% CI: 1.56–1.96) and double the likelihood of being prescribed psychotropic medication (2.17, 2.00–2.35) associated with having a positive COVID-19 test. The absolute risks were low: an estimated 1.4% of COVID positive patients presented with psychiatric morbidity at 6 months, compared to 0.9% of controls (Supplementary Table 5).

There was evidence of an increased risk in almost all outcomes considered. The largest increases were for receipt of antipsychotics (aHR: 6.94, 95%CI 4.32–11.15), fatigue (5.79, 5.05–6.61), receipt of non-benzodiazepine hypnotics (4.81, 3.78–6.12), receipt of mood stabilisers (3.47, 2.55–4.72), and sleep problems (3.09, 2.50–3.81).

Effect modification by age was evident (Table 3), such that the association between COVID-19 infection and psychiatric morbidity was greater for older age groups (e.g. aHR 80+ 4.00, 95%CI 2.28– 6.99 and 16-24: 1.21, 0.98–1.51). For fatigue and sleep disorder, the association was greatest for those aged 60-69 and remained elevated for all groups. For all outcomes, women with COVID-19 positive tests had a higher incidence than men, however the relative increase associated with a COVID-19 positive test was larger for men. There was no evident effect modification by deprivation quintile, and for sleep or psychiatric morbidity the association was similar over follow-up, although for fatigue the strength of effect was greatest in the first month.

**Table 2:** Comparison of incident outcomes between COVID-19 test positive patients and controls, matched on year of birth, gender and general practice.

| Outcome | Group | Events | Rate, per 1000 person years | Hazard Ratio (HR) |  |
| --- | --- | --- | --- | --- | --- |
|  |  |  |  | Unadjusted | Adjusted<sup>a</sup> |
| Any psychiatric morbidity | COVID-19 | 447 | 30.24 | 1.72 [1.54, 1.92] | 1.75 [1.56, 1.96] |
|  | Controls | 1045 | 17.39 | Ref | Ref |
| Depression | COVID-19 | 230 | 15.52 | 1.62 [1.39, 1.89] | 1.64 [1.40, 1.92] |
|  | Controls | 571 | 9.49 | Ref | Ref |
| Anxiety | COVID-19 | 298 | 20.12 | 1.77 [1.54, 2.03] | 1.85 [1.61, 2.13] |
|  | Controls | 671 | 11.15 | Ref | Ref |
| Psychosis | COVID-19 | 21 | 1.41 | 2.26 [1.31, 3.90] | 1.88 [0.89, 3.94] |
|  | Controls | 36 | 0.60 | Ref | Ref |
| Self-harm | COVID-19 | 13 | 0.87 | 2.08 [1.06, 4.07] | 2.05 [0.93, 4.55] |
|  | Controls | 25 | 0.41 | Ref | Ref |
| Sleep disorders | COVID-19 | 190 | 12.82 | 3.24 [2.67, 3.93] | 3.09 [2.50, 3.81] |
|  | Controls | 236 | 3.92 | Ref | Ref |
| Fatigue | COVID-19 | 579 | 39.29 | 5.87 [5.16, 6.68] | 5.79 [5.05, 6.63] |
|  | Controls | 414 | 6.88 | Ref | Ref |
| Any psychotropic medication | COVID-19 | 1055 | 72.05 | 2.33 [2.16, 2.52] | 2.17 [2.00, 2.35] |
|  | Controls | 1848 | 30.82 | Ref | Ref |
| Antidepressants | COVID-19 | 583 | 39.54 | 1.74 [1.58, 1.92] | 1.67 [1.51, 1.85] |
|  | Controls | 1358 | 22.62 | Ref | Ref |
| Benzodiazepines | COVID-19 | 266 | 17.95 | 3.63 [3.06, 4.30] | 3.30 [2.71, 4.01] |
|  | Controls | 305 | 5.07 | Ref | Ref |
| Nonbenzodiazepine hypnotics | COVID-19 | 173 | 11.67 | 4.87 [3.90, 6.09] | 4.81 [3.78, 6.12] |
|  | Controls | 148 | 2.46 | Ref | Ref |
| Antipsychotics | COVID-19 | 83 | 5.59 | 6.35 [4.45, 9.05] | 6.94 [4.32, 11.15] |
|  | Controls | 63 | 1.05 | Ref | Ref |
| Mood stabilisers | COVID-19 | 105 | 7.08 | 3.72 [2.85, 4.87] | 3.47 [2.55, 4.72] |
|  | Controls | 115 | 1.91 | Ref | Ref |

<sup>a</sup>Adjusted for ethnicity, smoking, BMI and comorbidities and after multiple imputation.

**Table 3:** Interactions between COVID-19 infection and covariates

|  | Psychiatric morbidity |  |  |  | Fatigue |  |  |  | Sleep |  |  |  |
| --- | --- | --- | --- | --- | --- | --- | --- | --- | --- | --- | --- | --- |
|  | Rate |  |  |  | Rate |  |  |  | Rate |  |  |  |
| Group | COVID-19 | Cont. | aHR<sup>a</sup> [95 % CI] | p-value<sup>b</sup> | COVID-19 | Cont. | aHR<sup>a</sup> [95 % CI] | p-value<sup>b</sup> | COVID-19 | Cont. | aHR<sup>a</sup> [95 % CI] | p-value<sup>b</sup> |
| Gender |  |  |  | 0.009 |  |  |  | 0.006 |  |  |  | 0.010 |
| Female | 37.11 | 24.06 | 1.55 [1.34, 1.80] |  | 43.01 | 8.68 | 5.02 [4.24, 5.94] |  | 8.31 | 3.50 | 2.28 [1.66, 3.12] |  |
| Male | 24.14 | 11.53 | 2.11 [1.76, 2.53] |  | 27.93 | 3.67 | 7.36 [5.90, 9.19] |  | 12.45 | 3.07 | 3.99 [3.00, 5.31] |  |
| Age-group |  |  |  | <0.001 |  |  |  | 0.0001 |  |  |  | 0.009 |
| 16-24 | 41.69 | 34.49 | 1.21 [0.98, 1.50] |  | 14.46 | 5.43 | 2.79 [1.93, 4.04] |  | 4.32 | 2.55 | 1.95 [1.09, 3.49] |  |
| 25-34 | 31.01 | 21.29 | 1.52 [1.16, 1.98] |  | 20.37 | 5.17 | 4.02 [2.81, 5.75] |  | 5.32 | 2.51 | 2.23 [1.25, 3.99] |  |
| 35-49 | 27.92 | 15.20 | 1.91 [1.52, 2.42] |  | 39.67 | 5.79 | 6.45 [5.03, 8.28] |  | 9.72 | 3.05 | 3.22 [2.16, 4.79] |  |
| 50-59 | 25.18 | 11.80 | 2.15 [1.61, 2.87] |  | 42.38 | 4.80 | 8.01 [5.96, 10.78] |  | 12.94 | 2.69 | 4.18 [2.68, 6.51] |  |
| 60-69 | 22.91 | 7.91 | 3.06 [1.98, 4.74] |  | 38.94 | 3.95 | 8.85 [5.84, 13.40] |  | 11.96 | 1.96 | 7.47 [3.81, 14.65] |  |
| 70-79 | 21.78 | 5.90 | 3.53 [1.80, 6.94] |  | 37.80 | 4.74 | 7.00 [4.10, 11.96] |  | 9.12 | 2.97 | 3.40 [1.37, 8.48] |  |
| 80+ | 46.06 | 11.97 | 4.00 [2.28, 6.99] |  | 32.17 | 9.76 | 5.03 [2.76, 9.17] |  | 11.50 | 5.88 | 1.17 [0.50, 2.74] |  |
| IMD quintile |  |  |  | 0.416 |  |  |  | 0.055 |  |  |  | 0.988 |
| Highest | 21.93 | 15.21 | 1.45 [1.01, 2.10] |  | 36.38 | 5.44 | 5.87 [4.14, 8.31] |  | 5.78 | 2.16 | 3.30 [1.73, 6.32] |  |
| 2<sup>nd</sup> | 33.39 | 18.14 | 1.79 [1.37, 2.34] |  | 43.24 | 4.51 | 8.66 [6.25, 12.00] |  | 7.21 | 2.45 | 3.31 [1.88, 5.83] |  |
| 3<sup>rd</sup> | 32.91 | 16.96 | 1.89 [1.42, 2.52] |  | 29.83 | 4.49 | 6.27 [4.41, 8.93] |  | 9.65 | 3.09 | 3.18 [1.93, 5.24] |  |
| 4<sup>th</sup> | 28.56 | 19.39 | 1.51 [1.18, 1.93] |  | 28.98 | 6.80 | 4.65 [3.45, 6.28] |  | 11.84 | 3.57 | 3.16 [2.06, 4.84] |  |
| Lowest | 33.02 | 16.61 | 1.99 [1.60, 2.48] |  | 32.45 | 5.82 | 5.00 [3.84, 6.51] |  | 8.82 | 3.31 | 2.81 [1.87, 4.22] |  |
| Follow-up time |  |  |  | 0.236 |  |  |  | <0.0001 |  |  |  | 0.12 |
| <1 month | 22.02 | 15.78 | 1.48 [1.23, 1.77] |  | 47.54 | 5.56 | 8.12 [6.71, 9.82] |  | 8.58 | 2.66 | 3.27 [2.33, 4.61] |  |
| 1-3 months | 31.58 | 17.69 | 1.86 [1.57, 2.21] |  | 24.66 | 5.64 | 4.47 [3.54, 5.63] |  | 8.75 | 3.57 | 2.27 [1.59, 3.24] |  |
| 3-6 months | 24.85 | 13.17 | 1.96 [1.48, 2.59] |  | 25.76 | 6.33 | 4.66 [3.35, 6.49] |  | 13.87 | 3.31 | 3.96 [2.49, 6.31] |  |
| 6 + months | 14.75 | 8.56 | 1.93 [1.14, 3.28] |  | 17.49 | 5.61 | 3.03 [1.77, 5.20] |  | 12.13 | 2.42 | 5.33 [2.40, 11.82] |  |

<sup>a</sup>Adjusted models included the covariates: ethnicity, comorbidities, smoking status, and BMI; <sup>b</sup>p-value testing for equivalence of hazard ratios

### Outcomes in those with prior history of common mental illness, psychosis, fatigue and sleep problems

For those with pre-existing common mental illness (depression or anxiety disorders), there was no increased risk associated with a positive COVID-19 test of subsequent depression or anxiety events (Figure 2, Supplementary Table 1). However, there was a small increase in the risk of new prescriptions for antidepressants (aHR 1.17, 1.10-1.25) and a larger increase for new prescriptions for benzodiazepines (1.84, 1.59-2.13). For those with depression or anxiety disorders, there was more than a doubling of the risk of subsequent fatigue associated with COVID-19 infection (2.27, 1.97-2.61) and a small increase in the risk of subsequent sleep problems (1.23, 1.04-1.46).

**Figure 2:**
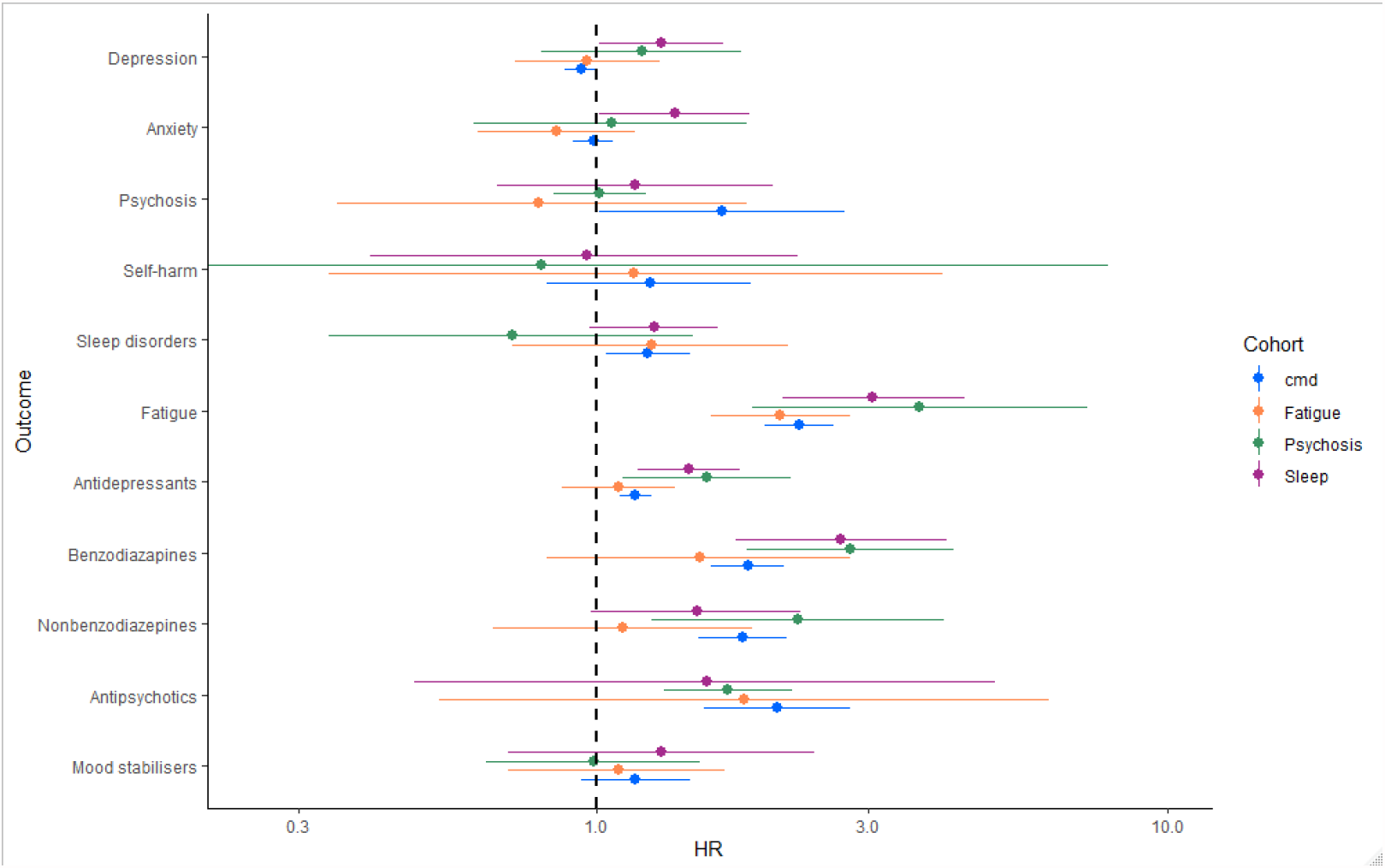
Association between testing positive for COVID-19 and psychiatric morbidity, sleep problems, fatigue and psychotropic medications (excluding those prescribed 6 months prior to index date), for those with pre-existing common mental illness, psychosis, fatigue or sleep problems

There was little evidence of an increased risk of depression or anxiety for those with pre-existing psychosis or fatigue; however, the confidence intervals were wide. There was evidence of an increased risk of depression and anxiety disorders associated with having a positive COVID-19 test for those with pre-existing sleep problems (aHR for depression 1.30, 1.01-1.67, for anxiety disorders 1.37, 1.01-1.86) and an increased risk of subsequent fatigue associated with having a positive COVID-19 test for all matched cohorts.

### Negative test and flu cohorts

There was a 1.57 times (1.51-1.63) increased risk of incident psychiatric morbidity for those with a negative COVID-19 test compared to comparators (Figure 3). There was a similar strength association to the main analysis between having a negative test and the risk of all subcategories of psychiatric morbidity and a weaker association with fatigue and sleep problems (Supplementary Table 2). There was a considerably greater association between having a flu-like illness and either incident psychiatric morbidity, sleep, fatigue or psychotropic prescribing than that seen in the main analysis, albeit with wide confidence intervals.

**Figure 3.**
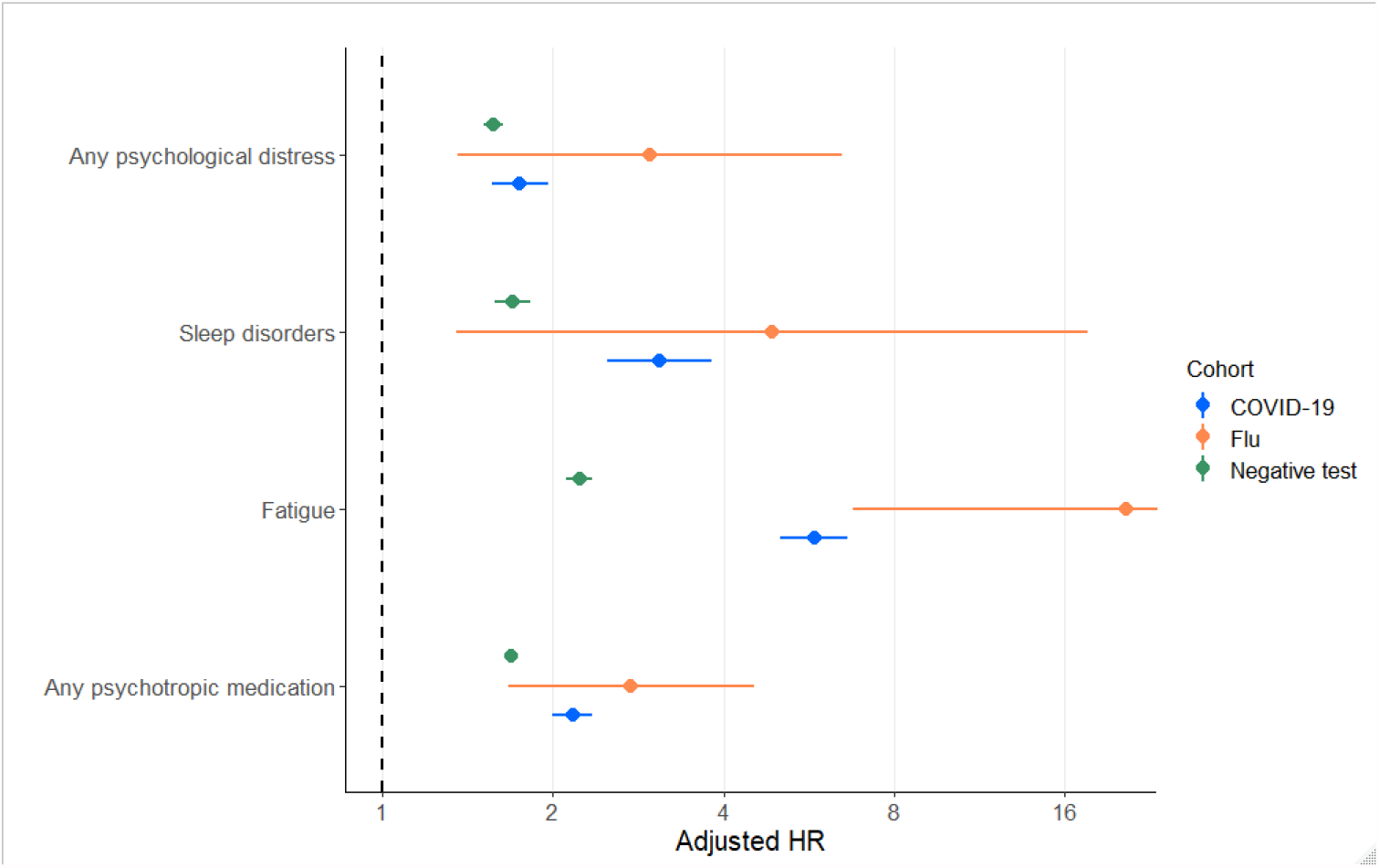
Adjusted hazard ratios from incident cohort, negative exposure cohort and flu cohort

### Sensitivity analyses

There were some significant interactions between period of positive test and the exposure. For individuals who tested positive prior to 1^st^ September 2020, there was a considerably stronger association between having a positive COVID-19 test and psychiatric morbidity or psychotropic medication than those who tested positive on or after 1^st^ September 2020 (Supplementary Table 3). There was little evidence that the estimate relating to fatigue or sleep problems was different between these two periods. The estimates, after adjusting for a propensity score, were of a similar magnitude to that seen in the main analysis (Supplementary Table 4).

## Discussion

In line with our primary hypothesis, we report that infection confirmed with a positive COVID-19 PCR test is associated with increased incidence of psychiatric morbidity, sleep problems and fatigue in the months that follow. In addition, there was a large increase in the rate of psychotropic prescribing, particularly for antipsychotic and benzodiazepine medications. Counter to our hypothesis, those with prior mental illness did not appear to have an elevated risk of subsequent psychiatric morbidity associated with having tested positive for COVID-19, although they were at elevated risk of fatigue or sleep problems. The relative increase in psychiatric morbidity, fatigue and sleep problems associated with COVID-19 infection was larger for older people and (counter to our initial hypothesis) men. The increased risk of fatigue was particularly evident in the first four weeks after an infection.

Additional analyses provide doubt about the degree to which COVID-19 infection is causing the increased risk of psychiatric morbidity. Notably, against matched comparators, those with a negative COVID-19 test, after adjustment for measured confounders, also saw a substantial increase in their risk of incident psychiatric morbidity. This association was of similar magnitude and with overlapping confidence intervals to that observed in people with a positive COVID-19 test. Having a negative test for SARS-CoV-2, in and of itself, is not likely to cause psychiatric morbidity through any direct mechanism. This ‘negative control’, therefore, reveals that individuals who attended for a test form a selected group with an underlying tendency, or higher risk of psychiatric morbidity during the pandemic; and this influence is not accounted for by measured variables included in the analysis. Several unobserved confounders might be influencing these results. For example, people who are health or care workers are more likely to come in to contact with the virus and require testing, and experience higher levels of psychiatric morbidity, particularly during the pandemic[16]. Also, those who seek a test might be experiencing health anxieties, that might indicate future mental illness needing treatment. These factors may have more influence on testing during the early stages of the pandemic, when testing was less widely available, and this may explain why the estimated increased in psychiatric morbidity associated with COVID-19 was stronger for those testing positive during the first wave.

We investigated those exposed to flu in order to examine the specificity of the relationship with COVID-19 over other respiratory diseases. However, for all outcomes, the increase in risk was considerably larger for attenders at general practice with flu than with COVID-19. This is likely because these form a particularly selective group: overall there were fewer cases of flu observed during the pandemic[17] and those who present may have been more likely to have significant morbidity and/or psychological vulnerabilities.

Our finding contrasts with two recent studies using US administrative data: these reported that individuals with positive tests for COVID-19 had roughly double the risk of subsequent psychiatric illness compared to those with influenza infection[5,6]. There are some key design differences that may explain this: the US analysis did not follow-up those with COVID-19 and the controls from the same date; nor did they adjust for geographical area. Because the pandemic has ecological effects on localised healthcare systems, as well as on population mental health, this could introduce substantive biases[18]. Also, the US may have confounding that is less evident in the UK. For example, in the US, areas with high levels of deprivation and inequality had significantly higher incidence of COVID-19[19,20], these areas are also likely to have higher rates of mental illness[21,22].

Our analysis is in agreement with a Danish registry study that did not find an association between testing positive for COVID-19, versus testing negative, and subsequent mental illness[7]. This analysis excluded those hospitalised for COVID-19 (which we were unable to do); however, we note that the US study reported similar estimates for those hospitalised and those not.

This research has far broader implications than improving methodological approaches. Our findings contribute to the current debate surrounding new resource allocation for unmet clinical need post pandemic[23]. The absence of data for a specific causal link between COVID-19 and subsequent psychiatric and associated sequelae, a better allocation of resource might be to expand existing services that already address post-viral fatigue. This would prevent the need for individuals to have had COVID-19 in order to access the service. This appears increasingly important if predictions of a particularly bad 2021-2022 flu season come to bear. Data from prior studies with random sampling suggests that anxiety and stress increased during the pandemic and some did not recover[24,25]; irrespective of COVID status, these individuals need adequate support taking account of the likely picture of physical and mental health symptoms.

Other designs may be more appropriate to investigate the specific effect of COVID-19 infection on mental health outcomes. For example, individuals, randomly selected, who took part in serological surveys might be more representative of the general population and, thus, less susceptible to bias. We conclude that the information to-date does not support a direct effect of COVID-19 and subsequent psychiatric morbidity and that more work is needed to understand if causal connections exist and what services might anticipate in relation to future increases in clinical need.

## Supporting information

RECORD

## Data Availability

Data sharing: Read codes used are published on https://Clinicalcodes.org. Electronic health records are, by definition, considered sensitive data in the UK by the Data Protection Act and cannot be shared via public deposition because of information governance restriction in place to protect patient confidentiality. Access to data are available only once approval has been obtained through the individual constituent entities controlling access to the data. The primary care data can be requested via application to the Clinical Practice Research Datalink (www.cprd.com/researcher), secondary care data can be requested via application to the hospital episode statistics from the UK Health and Social Care Information Centre (www.hscic.gov.uk/hesdata).

## Acknowledgments

This study was conducted using data from the CPRD obtained under licence from the UK Medicines and Healthcare products Regulatory Agency (MHRA). The data are provided by patients and collected by the NHS as part of their care and support. The interpretation and conclusions contained in this study are those of the authors alone, and not necessarily those of the MHRA, NHS, NIHR, or the UK Department of Health and Social Care. We would like to acknowledge all the data providers and general practices that made the anonymised data available for research. The study was approved by the Independent Scientific Advisory Committee for CPRD research (20_094R2).

