## Supplementary for "COVID-19 infection and subsequent psychiatric morbidity, sleep problems and fatigue: analysis of an English primary care cohort of 226,521 positive patients"

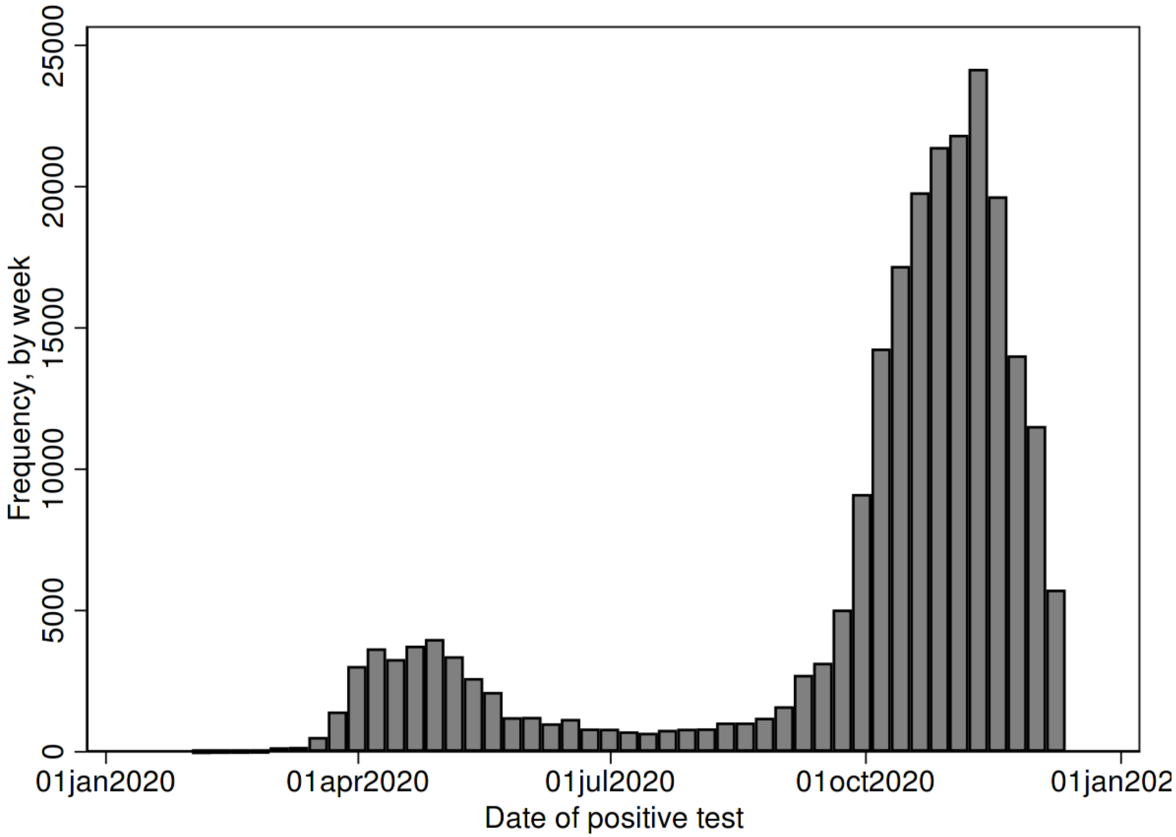


**Supplementary Figure 1: Histogram of frequency of positive COVID-19 test results by date during 2020**

**Supplementary Table 1: Psychiatric morbidity, sleep problems, fatigue and uniquely prescribed psychotropic medications for those with pre-existing common mental illness, psychosis, fatigue or sleep problems matched on year of birth, gender and general practice**

| | Common mental illness | | | Psychosis | | | Fatigue | | | Sleep | | |
|---|---|---|---|---|---|---|---|---|---|---|---|---|
| | Rate | | | Rate | | | Rate | | | Rate | | |
| Outcome | COVID-19 | Controls | aHR [95 % CI] | COVID-19 | Controls | aHR [95 % CI] | COVID-19 | Controls | aHR [95 % CI] | COVID-19 | Controls | aHR [95 % CI] |
| Depression | 352.47 | 379.84 | 0.94 [0.88, 1.00] | 155.32 | 138.13 | 1.20 [0.80, 1.79] | 148.96 | 167.23 | 0.96 [0.72, 1.29] | 172.16 | 132.86 | 1.30 [1.01, 1.67] |
| Anxiety | 263.13 | 280.83 | 0.99 [0.91, 1.07] | 77.92 | 94.09 | 1.06 [0.61, 1.83] | 131.00 | 144.98 | 0.85 [0.62, 1.17] | 137.98 | 94.97 | 1.37 [1.01, 1.86] |
| Psychosis | 24.92 | 24.64 | 1.66 [1.01, 2.72] | 623.40 | 639.56 | 1.01 [0.84, 1.22] | 10.12 | 13.32 | 0.79 [0.35, 1.83] | 19.31 | 17.32 | 1.17 [0.67, 2.04] |
| Self-harm | 15.77 | 11.63 | 1.24 [0.82, 1.87] | 13.41 | 20.29 | 0.80 [0.08, 7.90] | 5.05 | 4.61 | 1.16 [0.34, 4.04] | 8.11 | 6.91 | 0.96 [0.40, 2.26] |
| Sleep disorders | 65.88 | 51.79 | 1.23 [1.04, 1.46] | 54.19 | 60.76 | 0.71 [0.34, 1.48] | 48.43 | 37.56 | 1.25 [0.71, 2.17] | 123.79 | 94.83 | 1.26 [0.97, 1.63] |
| Fatigue | 105.61 | 44.47 | 2.27 [1.97, 2.61] | 95.76 | 32.38 | 3.69 [1.88, 7.26] | 153.38 | 84.15 | 2.10 [1.59, 2.78] | 92.03 | 36.68 | 3.05 [2.12, 4.41] |
| Antidepressants | 398.80 | 323.75 | 1.17 [1.10, 1.25] | 227.46 | 164.74 | 1.56 [1.11, 2.19] | 222.66 | 186.47 | 1.09 [0.87, 1.37] | 243.66 | 149.77 | 1.45 [1.18, 1.78] |
| Benzodiazepines | 108.91 | 52.94 | 1.84 [1.59, 2.13] | 225.51 | 79.37 | 2.78 [1.83, 4.22] | 67.53 | 33.46 | 1.52 [0.82, 2.79] | 87.46 | 32.78 | 2.68 [1.75, 4.12] |
| Non-benzodiazepine hypnotics | 64.04 | 38.97 | 1.80 [1.51, 2.16] | 88.59 | 56.92 | 2.25 [1.25, 4.06] | 38.16 | 27.24 | 1.11 [0.66, 1.88] | 63.71 | 39.40 | 1.50 [0.98, 2.28] |
| Antipsychotics | 46.35 | 22.99 | 2.07 [1.54, 2.78] | 381.26 | 252.79 | 1.70 [1.31, 2.20] | 20.26 | 10.76 | 1.81 [0.53, 6.22] | 39.81 | 20.78 | 1.56 [0.48, 5.00] |
| Mood stabilisers | 41.40 | 34.54 | 1.17 [0.94, 1.46] | 158.29 | 108.65 | 0.99 [0.64, 1.52] | 26.67 | 22.58 | 1.09 [0.70, 1.68] | 45.04 | 27.37 | 1.30 [0.70, 2.41] |

aHR = adjusted hazard ratios
Rate per 1,000 person years
Note: all medication outcomes refer to prescriptions not previously prescribed 6 months prior to index date

**Supplementary Table 2: Comparison of adjusted hazard ratios from matched positive, negative test and flu cohorts**

| | Matched cohort | | |
|---|---|---|---|
| **Outcome** | **Positive** | **Negative test** | **Flu** |
| Any psychological distress | 1.75 [1.56, 1.96] | 1.57 [1.51, 1.63] | 2.97 [1.36, 6.48] |
| Depression | 1.64 [1.40, 1.92] | 1.52 [1.44, 1.60] | 3.50 [1.06, 11.56] |
| Anxiety | 1.85 [1.61, 2.13] | 1.65 [1.57, 1.73] | 3.87 [1.54, 9.69] |
| Psychosis | 1.88 [0.89, 3.94] | 1.36 [1.06, 1.75] | * |
| Self-harm | 2.05 [0.93, 4.55] | 1.36 [1.09, 1.69] | * |
| Sleep disorders | 3.09 [2.50, 3.81] | 1.70 [1.58, 1.83] | 4.88 [1.35, 17.57] |
| Fatigue | 5.79 [5.05, 6.63] | 2.23 [2.11, 2.35] | 20.57 [6.78, 62.37] |
| Any psychotropic medication | 2.17 [2.00, 2.35] | 1.69 [1.64, 1.74] | 2.75 [1.67, 4.52] |
| Antidepressants | 1.67 [1.51, 1.85] | 1.59 [1.53, 1.65] | 2.84 [1.71, 4.72] |
| Benzodiazepines | 3.30 [2.71, 4.01] | 2.05 [1.91, 2.19] | 16.81 [1.68, 167.91] |
| Nonbenzodiazepine hypnotics | 4.81 [3.78, 6.12] | 1.90 [1.73, 2.09] | 1.64 [0.18, 14.72] |
| Antipsychotics | 6.94 [4.32, 11.15] | 2.11 [1.76, 2.52] | * |
| Mood stabilisers | 3.47 [2.55, 4.72] | 1.83 [1.64, 2.05] | 2.62 [0.31, 22.32] |

*Insufficient events (<5) to fit model

**Supplementary Table 3: Effect estimates for those with COVID-19 positive test in the first or second wave (cut-off: 1st September 2020)**

| Outcome | Period of infection | Rate exposed | Rate unexposed | aHR | p-value |
|---|---|---|---|---|---|
| Any psychological distress | 1st wave | 32.27 | 14.54 | 2.25 [1.84, 2.74] | <0.0001 |
| | 2nd wave | 26.30 | 19.45 | 1.36 [1.15, 1.59] | |
| Sleep disorders | 1st wave | 17.93 | 4.21 | 3.65 [2.63, 5.06] | 0.093 |
| | 2nd wave | 9.10 | 3.55 | 2.48 [1.81, 3.40] | |
| Fatigue | 1st wave | 40.78 | 7.18 | 5.72 [4.56, 7.17] | 0.447 |
| | 2nd wave | 31.91 | 6.30 | 5.10 [4.19, 6.20] | |
| Any psychotropic medication | 1st wave | 98.36 | 31.74 | 2.78 [2.45, 3.15] | <0.0001 |
| | 2nd wave | 43.66 | 29.84 | 1.44 [1.27, 1.64] | |

p-value testing for equivalence of adjusted hazard ratios between first and second wave

**Supplementary Table 4: comparison of effect estimates from main analysis with that calculated controlling for a propensity score**

| Outcome | Main analysis | Propensity score control |
|---|---|---|
| Any psychological distress | 1.64 [1.45, 1.86] | 1.60 [1.41, 1.81] |
| Sleep disorders | 2.98 [2.37, 3.74] | 2.82 [2.28, 3.49] |
| Fatigue | 5.35 [4.61, 6.22] | 5.18 [4.48. 5.99] |
| Any psychotropic medication | 1.99 [1.82, 2.18] | 1.95 [1.79, 2.13] |

**Supplementary Table 5: proportion of positive testers and controls with outcomes after 6 months within each matched cohort**

| | Matched cohort | | | | | | | | | |
|---|---|---|---|---|---|---|---|---|---|---|
| | Incident | | CMD | | Psychosis | | Fatigue | | Sleep | |
| **Outcome** | **COVID-19** | **Controls** | **COVID-19** | **Controls** | **COVID-19** | **Controls** | **COVID-19** | **Controls** | **COVID-19** | **Controls** |
| Any psychological distress | 1.44 | 0.85 | 20.26 | 20.18 | 30.23 | 31.42 | 9.71 | 7.65 | 10.13 | 9.29 |
| Depression | 0.85 | 0.47 | 14.51 | 14.59 | 6.99 | 6.13 | 6.84 | 5.04 | 6.55 | 6.19 |
| Anxiety | 0.91 | 0.53 | 10.70 | 11.21 | 3.51 | 3.78 | 5.79 | 3.86 | 5.64 | 5.68 |
| Psychosis | 0.07 | 0.03 | 1.37 | 1.04 | 24.50 | 26.07 | 0.70 | 0.78 | 0.57 | 0.59 |
| Self-harm | 0.05 | 0.02 | 0.68 | 0.55 | 0.72 | 0.95 | 0.43 | 0.24 | 0.10 | 0.24 |
| Fatigue | 1.37 | 0.36 | 4.87 | 2.17 | 4.93 | 1.93 | 4.19 | 1.71 | 7.18 | 4.16 |
| Sleep | 0.72 | 0.21 | 3.18 | 2.43 | 2.74 | 2.97 | 6.05 | 4.63 | 2.71 | 1.71 |
| Any psychotropic medication | 3.54 | 1.61 | 83.35 | 83.11 | 94.45 | 94.62 | 42.21 | 34.86 | 31.89 | 29.42 |
| Antidepressants | 2.09 | 1.14 | 79.55 | 80.23 | 49.98 | 50.41 | 33.45 | 29.32 | 28.72 | 25.91 |
| Benzodiazepines | 0.80 | 0.30 | 8.85 | 6.74 | 20.50 | 14.94 | 6.55 | 3.40 | 3.39 | 2.91 |
| Nonbenzodiazepine hypnotics | 0.52 | 0.15 | 5.64 | 4.99 | 11.04 | 10.01 | 5.45 | 3.84 | 3.00 | 2.31 |
| Antipsychotics | 0.20 | 0.06 | 5.65 | 5.35 | 72.75 | 75.07 | 4.17 | 3.42 | 1.72 | 1.59 |
| Mood stabilisers | 0.41 | 0.12 | 8.29 | 7.87 | 40.45 | 36.79 | 8.00 | 6.31 | 4.77 | 4.36 |

**Supplementary Table 6: description of eligible cohort according to test and flu status over follow-up**

| Characteristic | COVID positive only, N=159,155 | COVID negative only, N= 1,464,693 | COVID positive and negative, N= 67,366 | Flu, N= 36,913 | Not COVID positive negative or flu, N=10,194,978 |
|---|---|---|---|---|---|
| Gender | | | | | |
| Female | 84,367 (53.0) | 854,045 (58.3) | 42,614 (63.3) | 22,633 (61.3) | 5,007,361 (49.1) |
| Male | 74,788 (47.0) | 610,648 (41.7) | 24,752 (36.7) | 14,280 (38.7) | 5,187,617 (50.9) |
| Median age [IQR] | 42 [27, 57] | 40 [29, 55] | 42 [28, 57] | 48 [35, 61] | 45 [31, 62] |
| Ethnicity | | | | | |
| White | 103,992 (65.3) | 1,020,184 (69.7) | 45,000 (66.8) | 24,183 (65.5) | 6,505,441 (63.8) |
| Asian | 17,630 (11.1) | 105,892 (7.2) | 7,140 (10.6) | 4,792 (13.0) | 887,280 (8.7) |
| Black | 5,208 (3.3) | 44,017 (3.0) | 2,052 (3.1) | 1,994 (5.4) | 454,195 (4.5) |
| Mixed | 2,323 (1.5) | 20,710 (1.4) | 919 (1.4) | 637 (1.7) | 163,957 (1.6) |
| Other | 1,889 (1.2) | 14,181 (1.0) | 667 (1.0) | 558 (1.5) | 161,045 (1.6) |
| Missing | 28,113 (17.7) | 259,709 (17.7) | 11,588 (17.2) | 4,749 (12.9) | 2,023,060 (19.8) |
| Median BMI [IQR] | 26.4 [23.0-30.7] | 26.0 [22.6-30.1] | 26.2 [22.7-30.8] | 27.0 [23.4-31.6] | 25.8 [22.6-29.7] |
| Psychiatric illness in the last 5 years | | | | | |
| Depression | 23,924 (15.0) | 294,753 (20.1) | 13,460 (20.0) | 9,566 (25.9) | 1,298,455 (12.7) |
| Anxiety disorders | 19,502 (12.3) | 238,155 (16.3) | 10,573 (15.7) | 7,178 (19.5) | 1,027,132 (10.1) |
| Psychosis | 1,542 (1.0) | 18,989 (1.3) | 1,088 (1.6) | 708 (1.9) | 107,745 (1.1) |
| Eating disorder | 852 (0.5) | 8,963 (0.6) | 461 (0.7) | 285 (0.8) | 37,801 (0.4) |
| Personality disorder | 418 (0.3) | 7,387 (0.5) | 270 (0.4) | 358 (1.0) | 34,824 (0.3) |
| Self-harm | 1,825 (1.2) | 25,893 (1.8) | 1,051 (1.6) | 896 (2.4) | 111,136 (1.1) |
| Fatigue | 11,581 (7.3) | 119,886 (8.2) | 6,095 (9.1) | 4,662 (12.6) | 541,824 (5.3) |
| Sleep disorder | 10,737 (6.8) | 105,440 (7.2) | 5,338 (7.9) | 4,339 (11.8) | 560,433 (5.5) |
| Medication in the last 5 years | | | | | |
| Antidepressants | 40,539 (25.5) | 458,703 (31.3) | 22,414 (33.3) | 15,384 (41.7) | 2,209,707 (21.7) |
| Benzodiazepines | 12,286 (7.7) | 144,620 (9.9) | 7,395 (11.0) | 5,669 (15.4) | 677,395 (6.6) |
| Nonbenzodiazepine hypnotics | 7,598 (4.8) | 91,944 (6.3) | 4,426 (6.6) | 3,773 (10.2) | 447,420 (4.4) |
| Antipsychotics | 2,765 (1.7) | 35,933 (2.5) | 2,256 (3.4) | 1,352 (3.7) | 172,385 (1.7) |
| Mood stabilisers | 9,518 (6.0) | 93,785 (6.4) | 5,106 (7.6) | 4,152 (11.3) | 489,268 (4.8) |

*Note that if reported flu over follow-up then patient was in flu category, regardless of other exposures.*
